# Polycomb-associated and Trithorax-associated developmental conditions – phenotypic convergence and heterogeneity

**DOI:** 10.1101/2024.07.12.24310328

**Authors:** Alice Smail, Eema Jawahiri, Kate Baker

## Abstract

Polycomb group (PcG) and Trithorax group (TrxG) complexes represent two major components of the epigenetic machinery. This study aimed to delineate phenotypic similarities and differences across developmental conditions arising from rare variants in PcG and TrxG genes, using data-driven approaches.

462 patients with a PcG or TrxG-associated condition were identified in the DECIPHER dataset. We analysed Human Phenotype Ontology (HPO) data to identify phenotypes enriched in this group, in comparison to other monogenic conditions within DECIPHER. We then assessed phenotypic relationships between single gene diagnoses within the PcG and TrxG group, by applying semantic similarity analysis and hierarchical clustering. Finally, we analysed patient-level phenotypic heterogeneity in this group, irrespective of specific genetic diagnosis, by applying the same clustering approach. Collectively, PcG/TrxG diagnoses were associated with increased reporting of HPO terms relating to integument, growth, head & neck, limb and digestive abnormalities. Gene group analysis identified three multi-gene clusters differentiated by microcephaly, limb/digit dysmorphologies, growth abnormalities and atypical behavioural phenotypes. Patient-level analysis identified two large clusters differentiated by neurodevelopmental abnormalities and facial dysmorphologies respectively, as well as smaller clusters associated with more specific phenotypes including behavioural characteristics, eye abnormalities, growth abnormalities and skull dysmorphologies. Importantly, patient-level phenotypic clusters did not align with genetic diagnoses. Data-driven approaches can highlight pathway-level and gene-level phenotypic convergences, and individual-level phenotypic heterogeneities. Future studies are needed to understand the multi-level mechanisms contributing to both convergence and variability within this population, and to extend data collection and analyses to later-emerging health characteristics.

## Introduction

Chromatin remodelling is a fundamental epigenetic mechanism that orchestrates development and maintains the stability of differentiated tissues. Polycomb group (PcG) and Trithorax group (TrxG) multi-subunit complexes have key roles in chromatin remodelling, and act antagonistically to regulate chromatin accessibility by inducing the repression and activation of gene expression respectively ^1,2^. PcG complexes mediate transcriptional repression by promoting nucleosomal compaction, as well as deterring TrxG action ^1,3,4^. TrxG complexes (including COMPASS methyltransferases and BAF (SWI/SNF) remodelling complexes) maintain active gene expression and oppose PcG action by relaxing nucleosomal compaction ^1,3,4^. A tightly regulated balance of PcG and TrxG activity is thus necessary for the precise timing of gene expression during mammalian development; in particular, the PcG-TrxG system has a vital role in neurogenesis, by mediating the balance of neural progenitor cell differentiation and self-renewal ^5^.

Multiple developmental conditions arise from pathogenic variants in PcG and TrxG genes, forming a subset of Mendelian Disorders of Epigenetic Machinery (MDEMs) ^6^. Whilst individually rare, PcG and TrxG-related conditions collectively account for approximately 8% of monogenic developmental disorders in the DECIPHER database ^7^. These conditions are phenotypically heterogeneous, with variability within and across different genetic diagnoses. It has been suggested that while MDEMs are monogenic, they are conceptually similar to complex disorders, with epigenomic dysregulations contributing to an array of different phenotypes and severities via downstream pathways ^8^. Patients with different PcG and TrxG-related conditions can share overlapping phenotypic characteristics: for example, broad similarities can be observed between BAF- and COMPASS-complex associated conditions ^9-14^. It is plausible that phenotypic similarities across conditions are due to epigenomic dysregulation at shared genomic loci and at similar developmental stages, leading to downstream effects on gene expression that affect convergent developmental pathways. However, there are also phenotypic divergences within this group: for example, opposing features relating to growth have been observed amongst PcG-related conditions arising from variants in PRC1, PRC2 and PR-DUB complex genes ^2,5^. Phenotypic descriptions of each MDEM in isolation may not fully capture the extent of overlap or distinctiveness between conditions, and may underplay the extent of variability within each condition. A phenotypic evidence base that accounts for this complexity could improve post-diagnostic counselling and stimulate hypothesis driven, clinically relevant research.

The current paper reports data-driven analysis, aiming to describe the phenotypic commonalities and heterogeneities of PcG and TrxG-related conditions. Our first objective was to identify phenotypes that occur at elevated frequencies across the population of patients with PcG and TrxG-associated conditions, in comparison to other monogenic conditions that are present in the DECIPHER dataset. If phenotypic enrichments exist for this broad group of conditions, this reinforces the evidence for pathway-level convergence in developmental mechanisms and clinical needs. Our second objective was to map gene-level heterogeneity within PcG and TrxG-related conditions. To do this, we investigated whether there are phenotypic clusters of PcG and TrxG-associated conditions when patient data are grouped by affected gene. If gene-driven clusters are present, this supports the recognition of clinical syndromes aligning with variants across several specific genes, and may point to shared epigenomic dysregulations. Our third objective was to map patient-level heterogeneity to explore whether there are phenotypic clusters of patients with PcG and TrxG-associated conditions; the results of this final analysis may support or challenge the alignment between MDEM-associated phenotype co-occurrences and variants in specific genes.

## Materials and Methods

### Gene List Curation

Gene Ontology (GO) annotations were used to collate a comprehensive PcG and TrxG gene list (Supplementary Figure 1A), following a similar approach to Ciptasari and van Bokhoven ^15^. GO Cellular Component terms selected to represent PcG complexes were ‘PRC1 complex’ (GO:0035102), ‘ESC/E(Z) complex’ (synonymous with ‘PRC2 complex’; GO:0035098) and ‘PR-DUB complex’ (GO:0035517). To gather terms related to SWI/SNF and COMPASS complexes, several child terms of ‘SWI/SNF superfamily-type complex’ (GO:0070603) and three terms that correspond to the three subtypes of COMPASS complex were selected. After collating 114 genes using relevant GO terms, a further 21 genes that are likely to possess PcG/TrxG involvement were added based on literature reviews, and 6 genes that lack direct or specific PcG/TrxG involvement were removed (Supplementary Table 1). This resulted in a final list of 129 PcG/TrxG genes.

### Cohort and phenotypes curation

The DECIPHER database is a repository of genotypic and Human Phenotype Ontology (HPO) standardised phenotypic data deposited by clinical geneticists and laboratory scientists ^7,16^. The DECIPHER open access dataset was filtered using the pre-defined PcG/TrxG gene list, to identify all pathogenic or likely pathogenic sequence variants located in a PcG-related or TrxG-related gene (Supplementary Figure 1B). 506 variants that were present across 499 patients were identified; importantly, all patients with more than one variant had variants present in the same gene. Patients with 0 HPO terms were removed (n=37), resulting in a cohort of 462 patients with pathogenic or likely pathogenic variants across 38 PcG/TrxG genes. 99 patients had PcG variants, 359 patients had TrxG variants, and 4 patients had variants in HCFC1 which has roles in both PcG (PR-DUB accessory subunit) and TrxG (SET1A/B-COMPASS complex subunit). All patients with autosomal variants were heterozygous (n=447), while all patients with X-linked variants were hemizygous (n=15). The majority (81%) of patients in this cohort had a de novo variant, while 13% had unknown inheritance and 6% had a maternally or paternally inherited variant. To obtain a comparison cohort, the remainder of the DECIPHER dataset was filtered to include unique patients with pathogenic or likely pathogenic sequence variants, and patients with 0 HPO terms were removed (n=5070).

### Group-level HPO enrichment analysis

To address the first study objective, the HPO terms reported for each patient were propagated using the Python package PyHPO, so that broader ‘top-level/parent’ phenotypes could be compared across groups, in addition to more specific ‘lower-level/child’ phenotypes ^16^. The median number of reported HPO terms, and top-level HPO terms for each patient in the PcG/TrxG and comparison cohorts were compared via Mann-Whitney U rank tests. We then compared the frequency of propagated HPO terms between the PcG/TrxG and comparison cohorts, using a two-proportions z-test with Benjamini-Hochberg correction for multiple testing (Figure 1a). First, we examined whether any top-level HPO terms were enriched in the PcG/TrxG population: for this analysis the top-level term ‘Abnormality of the musculoskeletal system’ was split into the descendent terms ‘Abnormality of the skeletal system’, ‘Abnormality of the musculature’ and ‘Abnormality of connective tissue’, which resulted in a total of 25 top-level HPO terms (Supplementary Table 2). Second, we examined lower-level term enrichments.

**Figure 1.**
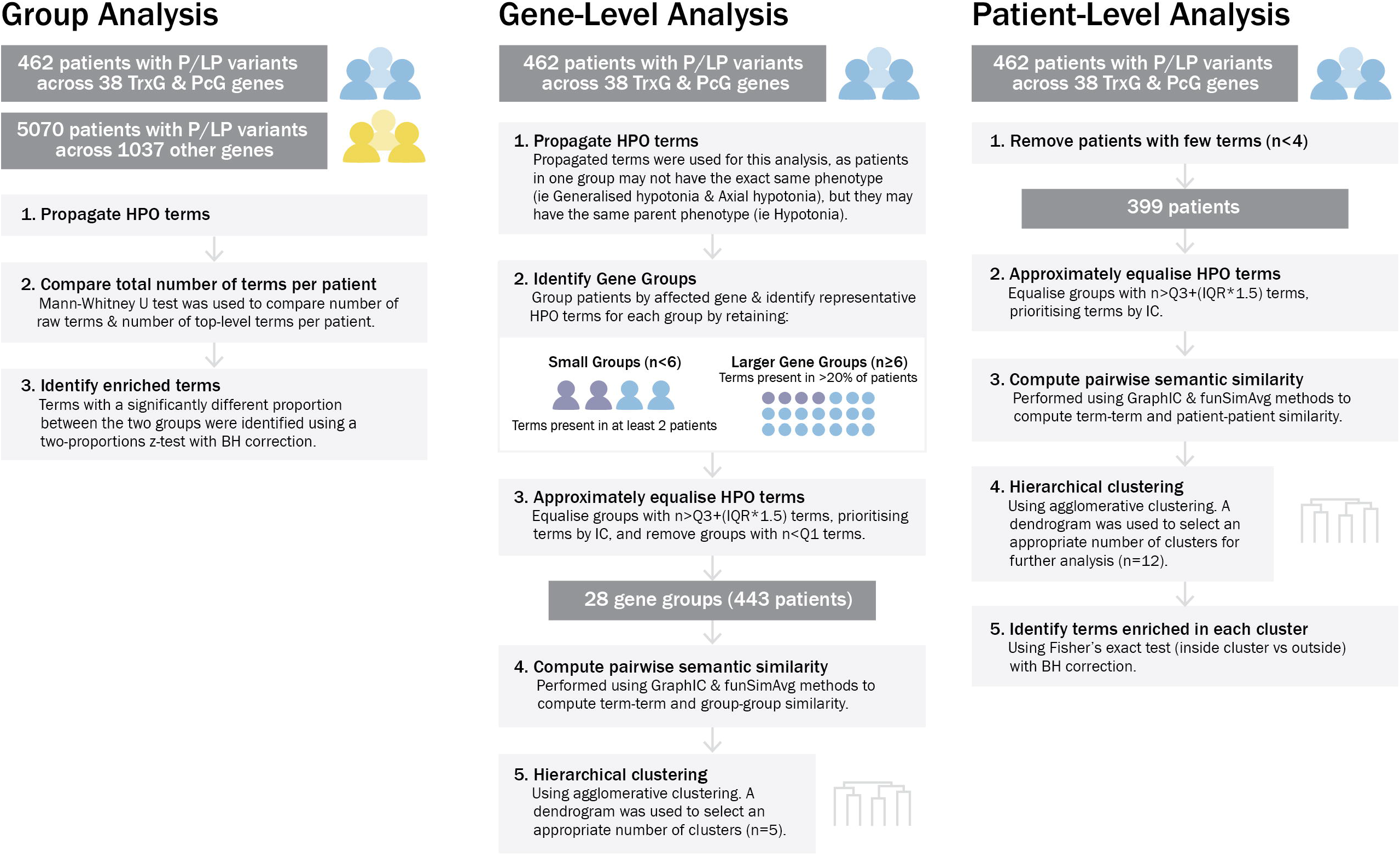
Overview of analysis methods.

### Gene-level HPO analysis

Figure 1b details the approach taken to examine gene-associated phenotypes within PcG- and TrxG-related conditions. This involved grouping patients by genetic condition, identifying a set of representative HPO terms of each gene group, and computing semantic similarity scores, before performing cluster analysis to identify phenotypic similarity between gene groups. To identify ‘representative’ HPO terms for each gene group, unique propagated HPO terms were collated, and depending on gene group size, HPO terms present in at least 2 patients or 20% of patients were retained (Supplementary Table 3). At this stage, each gene group contained a variable number of HPO terms, with some groups containing higher or lower number of terms. This could impact the cluster analysis outcomes by biasing the gene groups with similar numbers of HPO terms to artefactually cluster together. To reduce this bias, the number of HPO terms per group were approximately equalised. Based on the interquartile range of the number of terms across all gene groups, the cut off for the minimum and maximum number of terms retained per gene group were determined (i.e., removing outliers). The HPO terms retained within the set limit were prioritised based on their information content (IC) ^17,18^. The IC of HPO terms is a measure of the rarity/ specificity of a term within a database, where the IC of term t is given by, IC(t) = -log p(t), and p(t) is the probability of occurrence of t. Thus, terms with higher IC reflect more specific and clinically informative terms. As such, in this analysis, terms with higher IC were prioritised to be retained within the IQR set limit of number of terms.

Next, the semantic similarity scores and pairwise gene group similarity scores for each pair of gene groups was computed, using the graph IC method followed by the funSimAvg method ^18^. This approach takes into account all common ancestors of pairs of terms. The resulting similarity matrix was used as input for clustering. The method of cluster analysis chosen for this study was agglomerative hierarchical clustering using complete linkage; hierarchical clustering does not require a pre-specified number of clusters, which is useful for this study, as the number of different phenotypic profiles across the cohort was not known before clustering. Clustering was carried out using scikit-learn and SciPy packages in Python, and number of clusters was selected via visual inspection of the dendrogram^19^. To illustrate phenotype sharing between genes which leads to clustering, networks of semantic similarity were visualised using Cytoscape. The method used to select HPO terms for network visualisation is detailed in Supplementary Table 4: in brief, terms were included if present in at least 4 gene groups, terms with listed descendent terms were excluded, and some terms were merged to provide clearer visualisation.

### Patient-level HPO analysis

Figure 1c outlines the second approach for identifying phenotypic clusters, which involved directly computing the semantic similarity between patients, prior to performing hierarchical clustering. Raw HPO terms associated with each patient were collated. Modifier terms as well as terms with descendent terms were removed (i.e. parent term was removed if its child term is present). Similar to the gene-level pre-processing approach, the number of HPO terms associated with each patient were approximately equalised by identifying the upper outlier term frequency (Q3 + 1.5 x IQR) across all patients and using this as an upper limit with terms prioritised by highest IC. The semantic similarity between each pair of patients was then computed, before performing hierarchical clustering as described previously. Following clustering, HPO terms associated with each patient were propagated and the proportion of patients with each term in each cluster was compared to the proportion of patients with each term outside of each cluster, using Fisher’s exact test with a Benjamini-Hochberg correction. HPO terms that were significantly enriched (p_adj_<0.05) in each cluster were identified.

## Results

### Group-level HPO enrichment analysis

The number of HPO terms per patient for the PcG- and TrxG-associated conditions and comparison cohorts are displayed in Supplementary Figure 2. There is a significant distribution shift (p=1.51e-10) towards a higher number of reported HPO terms for patients with PcG- and TrxG-associated conditions (median = 7) relative to the remainder of the DECIPHER dataset (median = 6). Similarly, using top-level HPO terms as a proxy for the number of organ systems affected, there is a significant shift (p=1.51e-13) towards a higher number of top-level HPO terms across patients with PcG- and TrxG-associated conditions (median = 5) relative to the remainder of the DECIPHER dataset (median = 4). Together, these findings indicate an increased number of phenotypes per patient, and a larger breadth of affected organs/organ systems, in patients with PcG- and TrxG-associated conditions compared to other monogenic conditions.

Using propagated terms for each patient, the percentage occurrence of top-level HPO terms among patients with PcG and TrxG-associated conditions was compared to the percentage occurrence of each term across the remainder of the DECIPHER dataset. Five top-level terms were significantly increased at p_adj_<0.005 among patients with PcG and TrxG-associated conditions (Figure 2, Supplementary Table 5): ‘Abnormality of the integument’ (p_adj_=2.04e-09), ‘Growth abnormality’ (p_adj_= 5.44e-08), ‘Abnormality of head or neck’ (p_adj_=1.33e-06) ‘Abnormality of the digestive system’ (p_adj_= 4.04e-06), ‘Abnormality of limbs’ (p_adj_=0.0012). ‘Abnormality of the nervous system’ was also marginally increased at p_adj_=0.019. No top-level terms were significantly reduced in the PcG/TrxG group. The distribution of each top-level term across each genetic condition is illustrated in Supplementary Figure 4; it can be observed that a substantial proportion of patients with each PcG and TrxG-associated condition have abnormalities of the head or neck, integument, limbs and nervous system, while growth abnormalities and digestive system abnormalities are more unevenly distributed.

**Figure 2.**
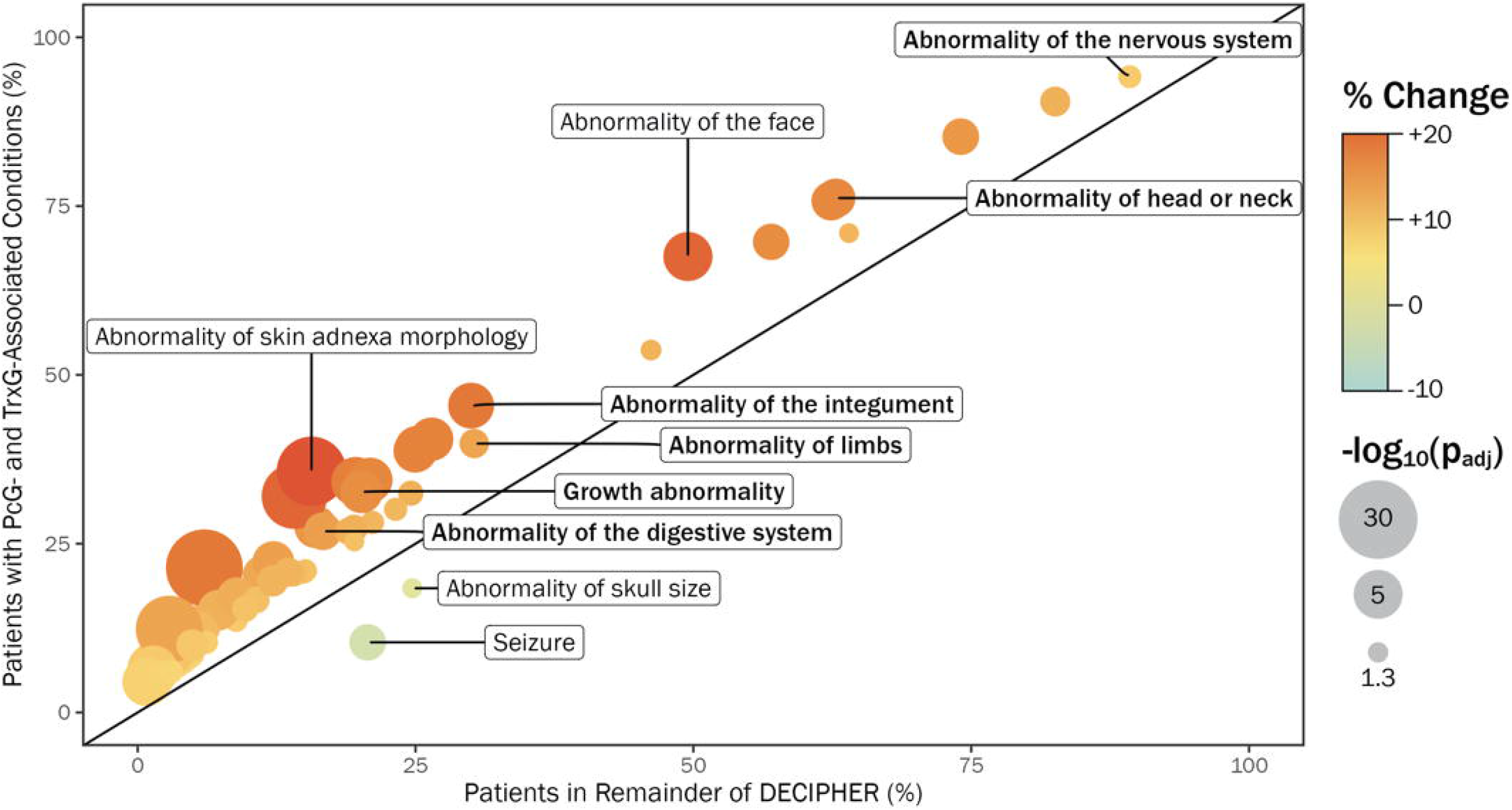
Group-level HPO analysis results. The proportion of each enriched (p_adj_<0.05) propagated HPO term among patients with PcG and TrxG-associated conditions is indicated by the y-axis, relative to the remainder of DECIPHER (x-axis). Terms with at least a ±3% change between the PcG/TrxG group and remainder of DECIPHER are shown. Significantly enriched top-level terms are labelled in bold; also labelled are the two terms with the largest positive percentage difference and the two terms with the largest negative difference between the groups.

Supplementary Table 6 lists specific HPO terms which were significantly increased or decreased at p_adj_<0.05, and that have a percentage difference of at least ±3% between the PcG/TrxG group and comparison group. 66 descendent terms of the 6 significantly enriched top-level terms were also significantly enriched in the PcG/TrxG group, and 2 descendent terms were significantly reduced (‘Seizure’ and ‘Abnormality of skull size’; Figure 2). 5 additional terms not descended from enriched top-level terms were significantly increased in the PcG/TrxG group. In summary, despite the genetic heterogeneity of PcG-related and TrxG-related conditions, there are detectable differences between this amalgamated population and the broader developmental disorders population: we observed differences in the median number of reported phenotypes, and differences across broad and specific phenotype frequencies.

### Gene-level HPO analysis: semantic similarity, clustering and network analysis

Representative HPO terms for each genetic diagnosis are listed in Supplementary Table 3. The computed semantic similarity matrix was used to perform hierarchical clustering (Figure 3A), resulting in three multi-gene clusters and 2 stand-alone gene groups. We examined clusters for potential bias arising from HPO term frequencies across each cluster (Supplementary Figure 3). Network analysis was applied to visualise the broad shared phenotypic characteristics of each cluster – for visualisation purposes, networks are presented separately for “Abnormalities of the nervous system” (Figure 3B) and descendent terms of “Head and neck abnormality”, “Abnormality of limbs”, “Abnormality of the integument” and “Growth abnormality” (Figure 3C). Broadly, it can be observed that genes within cluster 1 share growth abnormalities (including terms relating to overgrowth and restricted growth) and atypical behaviour; genes within cluster 3 share abnormalities of brain morphology, particularly microcephaly; genes within clusters 2 and 3 share limb abnormalities and facial dysmorphology. There was no clear pattern of PcG/TrxG complex membership aligning with phenotypic cluster membership i.e. COMPASS and PcG associated genes were distributed across all three clusters, while BAF complex genes were clustered into clusters 2 and 3.

**Figure 3.**
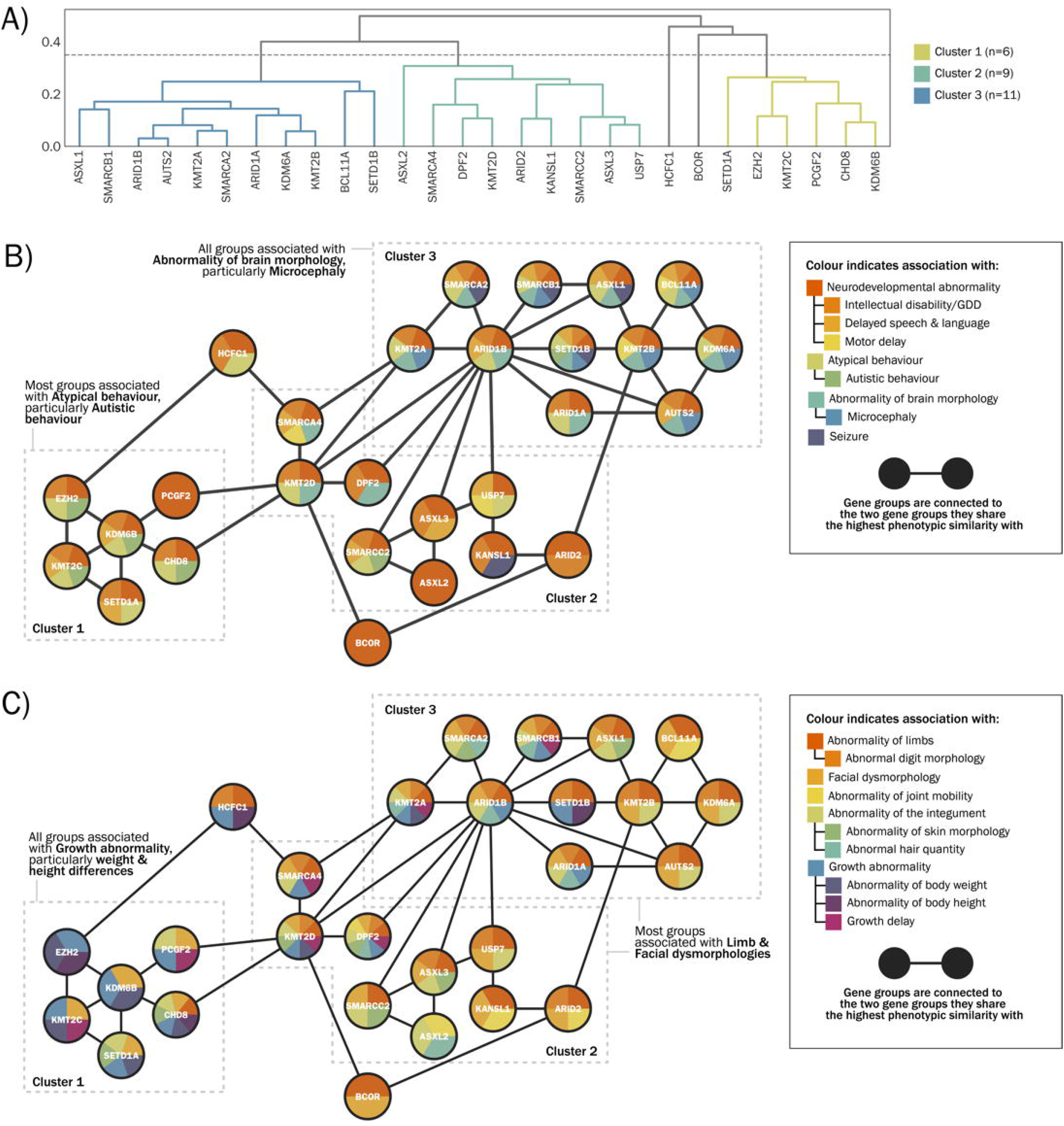
Gene-level clustering results. A) Dendrogram of gene-level clustering output. B) Network illustration of gene relationships within clusters, based on nervous system abnormalities present in at least 4 gene groups. C) Network illustration of gene relationships within clusters, based on abnormalities of physical development present in at least 4 gene groups.

### Patient-level HPO analysis: phenotypic similarity, clustering and cluster comparisons

To identify clusters of patients with phenotypic similarity within the PcG- and TrxG-associated conditions population, irrespective of specific genetic diagnosis, hierarchical clustering was performed on the unpropagated HPO terms for each individual patient. Following inspection of the dendrogram (Figure 4A), the population of patients was divided into 12 clusters, varying in size between 3 and 111 patients; this number of clusters was selected as it provides good insight into sub-clusters across the population, without separating larger clusters into smaller very similar clusters. Supplementary Figure 3 shows the HPO term numbers across each cluster. The phenotypic profile of each of the 12 clusters was then analysed using propagated patient-level terms, with significantly enriched terms identified at p_adj_<0.05 (Supplementary Table 7). Figure 4B summarises the two most significant HPO terms for each cluster. Cluster 1 contains the largest number of patients (n=111) and is enriched for a collection of neurodevelopmental abnormalities, particularly neurological speech impairment, in addition to abnormal muscle tone. Cluster 2 (n=91) is characterised by various abnormal facial morphologies, and Cluster 3 (n=48) has a higher proportion of patients with behavioural characteristics, including autistic behaviour. Cluster 4 (n=48) is characterised by abnormalities of the eye, while Cluster 5 (n=32) contains patients with growth abnormalities – both undergrowth (short stature, growth delay) and overgrowth (tall stature, overgrowth). Cluster 6 (n=30) is associated with skull dysmorphologies, particularly microcephaly. The remaining clusters contained 10 or fewer patients and were each defined by more specific HPO terms.

**Figure 4.**
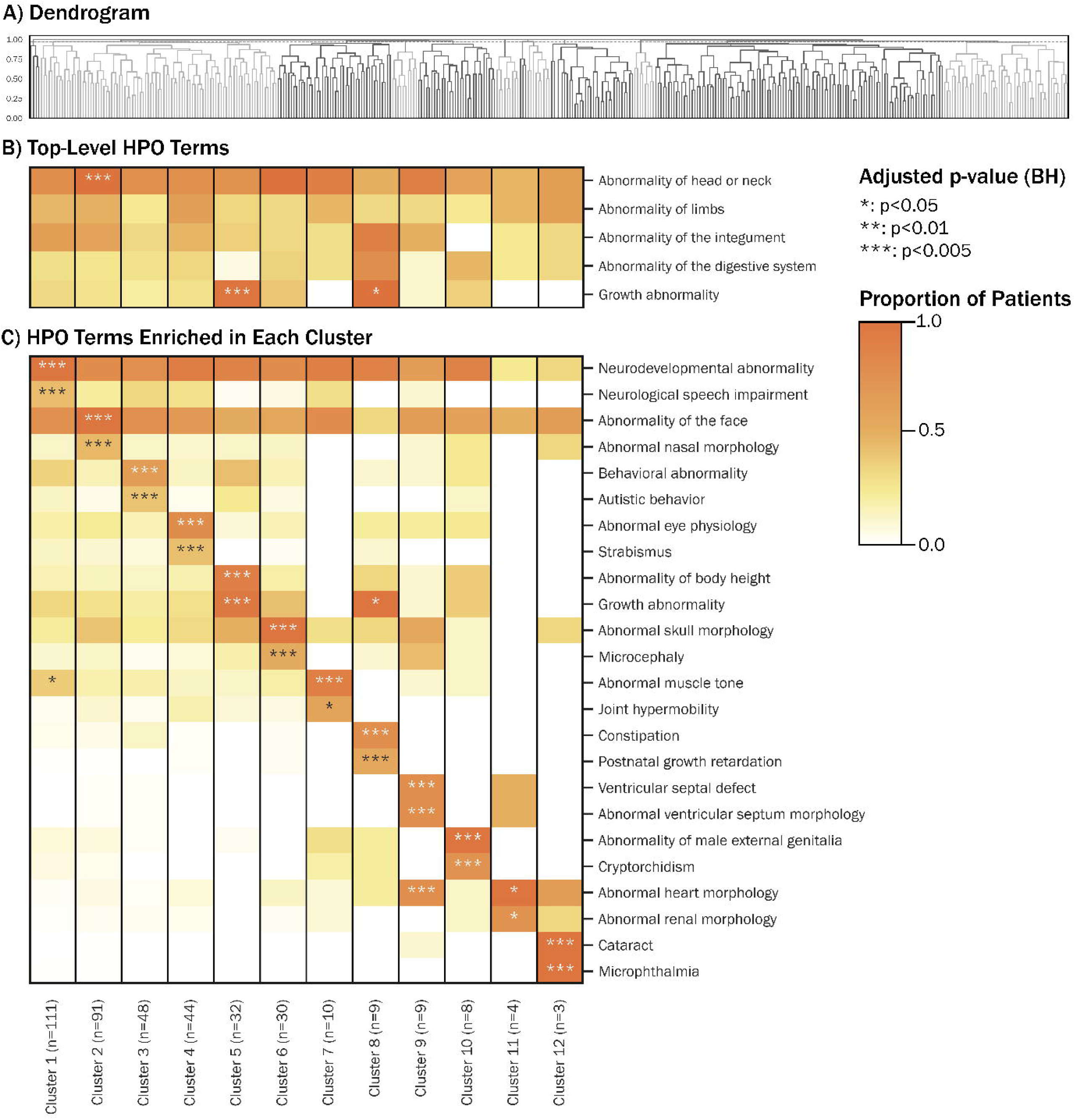
Patient-level clustering results. A) Patient-level dendrogram. B) Within-cluster proportional occurrence of top-level terms that were increased across the population as a whole. C) Within-cluster proportional occurrence of 2 representative HPO terms for each of the 12 clusters, which were identified using Fisher’s exact test.

Supplementary Figure 5 illustrates the distribution of cluster memberships for patients within each PcG- and TrxG gene diagnosis group. It can be observed that the majority of gene groups are phenotypically heterogeneous, and that phenotype clusters are distributed across genes. There are some exceptions, namely CHD8 (>50% patients within Cluster 11), SETD1A (>50% patients within Cluster 1) and BCOR (>50% patients within Cluster 12). However the absolute number of patients with each of these genetic diagnoses is low, hence this variation in proportional cluster membership may occur by chance.

## Discussion

This paper presents data-driven analyses of the phenotypic landscape of PcG- and TrxG-associated conditions. These analyses provide important insights into clinical convergences and divergences within this population, relevant to diagnosis, post-diagnostic counselling and management. Moreover, this study highlights opportunities and limitations of HPO computational methods for investigating disorder-associated pathways defined by convergent gene function.

We first assessed group-level differences in HPO term frequencies between PcG/TrxG-associated conditions and other monogenic developmental disorders. PcG/TrxG-associated conditions shared multi-system characteristics, with significantly increased frequency of abnormalities of growth, limbs, digestive system, integument and head or neck. More specific HPO terms reported in >20% of PcG/TrxG individuals include abnormal skin or hair morphology, abnormal ocular or oral morphology, and abnormal hands or upper limbs. These significant enrichments are useful phenotypic markers for confirming pathogenicity of PcG/TrxG variants. On the other hand, occurrences of each specific characteristic affect a minority of the population, emphasising the variability of this population, and need to assess combinations of phenotypes rather than each in isolation.

94% of patients with PcG/TrxG variants are reported to have nervous system abnormality, a modest elevation compared to other conditions (89%). ‘Global developmental delay’ (54% cases, 46% controls) and ‘Neurological speech impairment’ (25% cases, 19% controls) were also enriched, but terms relating to severity of developmental delay did not differ between groups. However, the reporting of specific cognitive and behavioural characteristics is sparse within DECIPHER, and HPO terminology is not designed for quantification of neurodevelopmental differences. ‘Abnormal cerebral white matter morphology’ is an enriched phenotype within the case group highlighting convergence on structural brain connectivity; seizures are reported at lower frequency (10% cases, 20% controls). Given the prevalence and variability of neurodevelopmental phenotypes within the PcG/TrxG group, investigation of multi-level mechanisms linking epigenetic regulation to brain development and cognitive vulnerabilities are warranted.

Building on group-level analysis of phenotype enrichments, our next goal was to assess phenotypic variability within PcG/TrxG-related conditions. We observed three clusters of genes which share broad top-level terms but differ in frequency of lower-level terms. Genes within clusters show multi-phenotype similarity to their within-cluster neighbours, but sparse connectivity to genes in other clusters. The phenotypes driving cluster separation within this dataset are likely to be partially influenced by the reporting biases of DECIPHER. However, these results are potentially hypothesis-setting for mechanistic studies: for example, the high phenotypic similarity between conditions associated with BAF, COMPASS and PcG subunits may reflect convergent impacts on gene expression^8^. However we also observed contrasting phenotypic profiles associated with functionally similar genes (for example, KDM6A and KDM6B); these differences may be influenced by variant-specific, cell-specific or timing-specific effects.

The pooled phenotypic profile for each gene does not take into account patient-level heterogeneity^20^. Therefore, we carried out a second clustering analysis including all patients with at least 4 reported HPO terms. We identified 12 phenotypically-defined clusters, which do not align with genetic diagnoses. Smaller clusters are driven by low prevalence phenotypes, whereas larger clusters are characterised by broad and frequent terms, with more subtle between-cluster differences. Phenotype cluster memberships may be influenced by unanalysed factors, such as the age of patients at phenotyping, and variable reporting by clinical geneticists, developmental paediatricians or neurologists. Importantly, gene groups were distributed across multiple clusters, and the majority of clusters were genetically heterogeneous, suggesting that phenotyping is not overly influenced by prior expectation of gene-specific features.

These results reinforce the complexity of MDEMs and the potential for early developmental mechanisms which cascade to system-level consequences that are not predictable by gene alone. However, limitations of our analyses include variable patient numbers across MDEMs in DECIPHER (more than 50% of the total group have one of four diagnoses), making it difficult to gauge and compare the phenotypic spectrum of every condition. There are disparities in phenotyping detail across the DECIPHER dataset - while some patients have an extensive list of fine-grained HPO terms, others have a single broad term, such as ‘Global developmental delay’; this may reflect the reality of a patient’s phenotype, or variations in phenotyping standards. However, we mitigated this bias by prioritising terms with higher IC (and therefore specificity), and application of a term equalisation step in our pre-processing to mitigate potential biases arising from disparities in HPO term numbers. Additionally, facial phenotypes and other specific morphological features can differentiate disorders from each other^21^. The fact that not every patient has a comprehensive list of specific phenotypes means that subtle dysmorphic differences will not contribute to our clustering analyses.

Our analyses focused on a subset of MDEMs, and future studies may be more informative by taking a more inclusive strategy to gene selection; we focused on the well-defined PcG and TrxG-associated complexes, while there is uncertainty as to what constitutes an ‘epigenetic’ protein involved indirectly in chromatin structural regulation. Despite this, there remains some ambiguity in our defined gene list: for example, while USP7 interacts with PRC1 variants, it may not strictly be considered a PcG protein^22^. Several genes have additional roles beyond the PcG-TrxG system, for example KDM2B functions as a demethylase at several histone sites, as well as mediating ubiquitination ^23^. There are also limitations arising from the classification of variants: this study relies on clinical reporting, and variants that may only partially explain phenotype were included. Additionally, this study did not take into account variant consequence; gain-of-function and loss-of-function variants may have different phenotypic consequences^24,25^.

One motivation for elucidating convergent clinical characteristics is that emerging treatment strategies may apply to a larger cohort beyond single genes. Ciptasari and van Bokhoven suggest that transcriptomic convergences across different MDEMs could be harnessed to develop effective interventions^15^. Drugs targeting epigenetic mechanisms or downstream gene expression have the potential to be efficacious in treating MDEMs^6^. HDAC inhibitors normalised H3K4 methylation and neurogenesis in a mouse model of Kabuki Syndrome^26^. CRISPR-based methods may also be applicable to MDEMs^27^. However, a major challenge is that therapies need to be administered at an appropriate developmental stage. Epigenetic interventions and CRISPR-based therapies risk introducing off-target changes^6^. The current study indicates that patient-relevant symptom domains are highly variable, hence target outcomes and therapeutic strategies will need to weigh-up potential benefits and risks on an individual basis.

The data-driven approach of this paper lays the foundation for future work disentagingly MDEM-phenotype relationships at scale. Future phenotyping research and clinical initiatives should move beyond congenital characteristics to encompass health-related outcomes across the lifespan.

## Data Availability

The dataset analysed during the current study is available in the DECIPHER open access database https://www.deciphergenomics.org/

## Code Availability

Code used for this project is available at github.com/alicesmail12/HPOAnalysis.

## Acknowledgments

This work was supported by UKRI/MRC funding (grant number MC_UU_00030/3). This study makes use of data generated by the DECIPHER community. A full list of centres who contributed to the generation of the data is available from https://deciphergenomics.org/about/stats and via email from. DECIPHER is hosted by EMBL-EBI and funding for the DECIPHER project was provided by the Wellcome Trust [grant number WT223718/Z/21/Z]. Those who carried out the original analysis and collection of the data bear no responsibility for the further analysis or interpretation of the data.

## Author Contribution Statement

AS and KB conceived the study. AS carried out data curation and analysis. EJ provided methodological advice. KB and AS wrote the manuscript, and all authors reviewed and approved the final version.

## Ethical Approval

The ethical framework for DECIPHER is provided at https://www.deciphergenomics.org/files/pdfs/decipher_ethical_framework.pdf

## Competing Interests

Nil to declare

## Notes

Funding support – UKRI/MRC (MC_UU_00030/3 to KB)

### Competing Interest Statement

The authors have declared no competing interest.

### Funding Statement

This study was funded by UKRI/MRC (MC_UU_00030/3)

### Author Declarations

The study used ONLY openly available human data that were originally located at www.deciphergenomics.org

