## Supplementary Figure for "Polycomb-associated and Trithorax-associated developmental conditions – phenotypic convergence and heterogeneity"

**Supplementary Figure 1**

**Gene list curation and cohort curation**

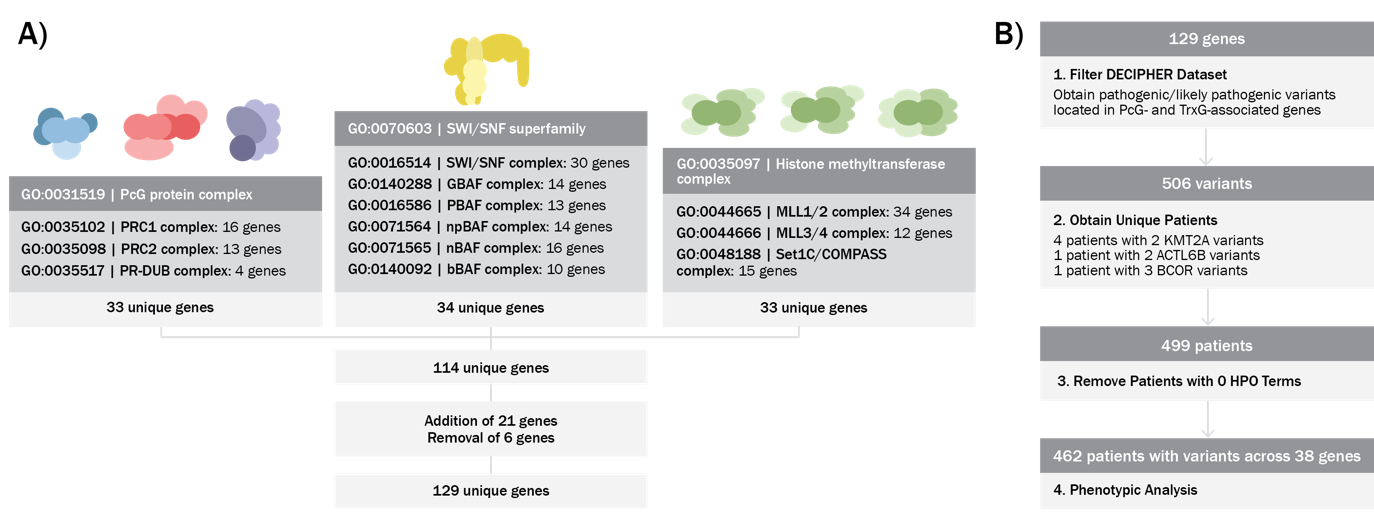

A) PcG / TrxG gene curation

B) DECIPHER dataset filtering methods

**Supplementary Figure 2**

**Distributions of total HPO term numbers in PcG/TrxG and comparison cohorts**

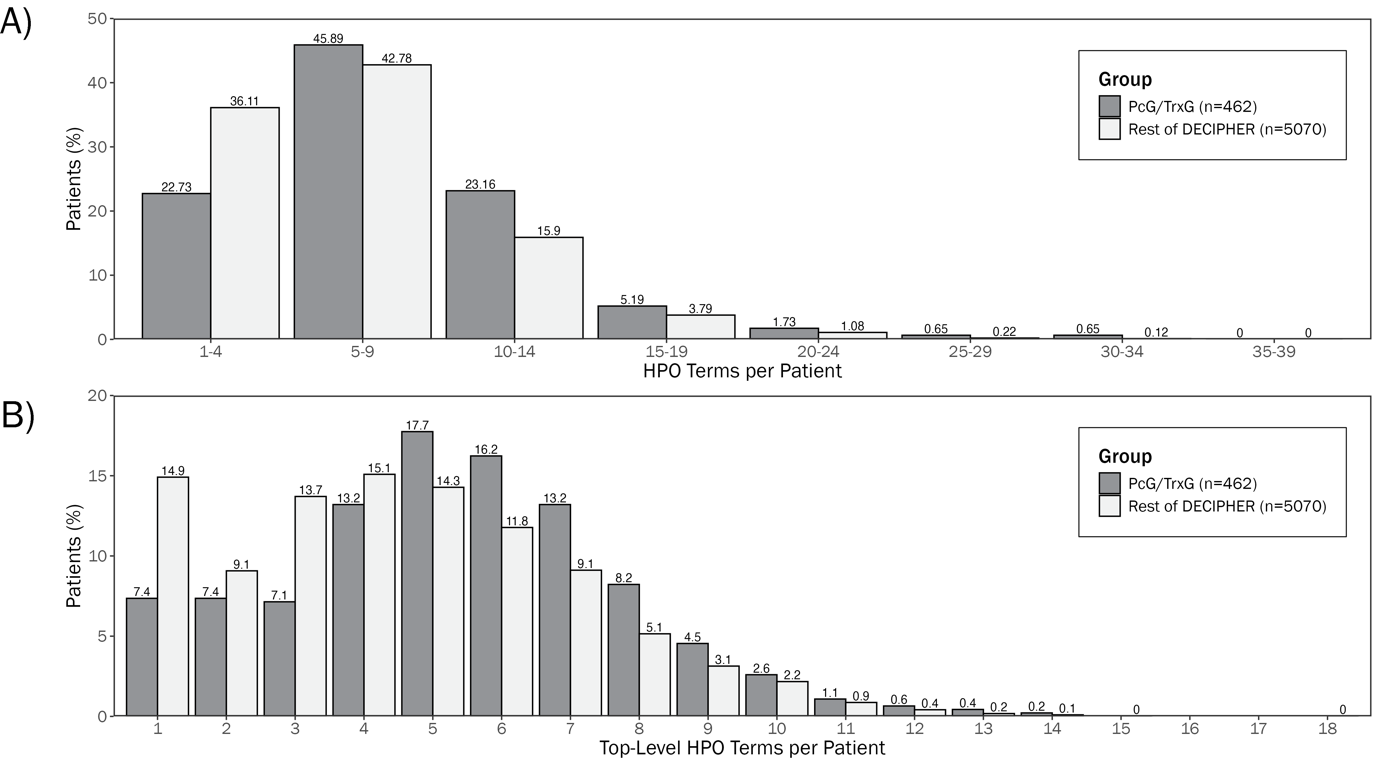

A) Number of raw HPO Terms per patient.

B) Number of top-level terms per patient after propagation.

**Supplementary Figure 3**

**HPO term list sizes for gene-level and patient-level clusters**

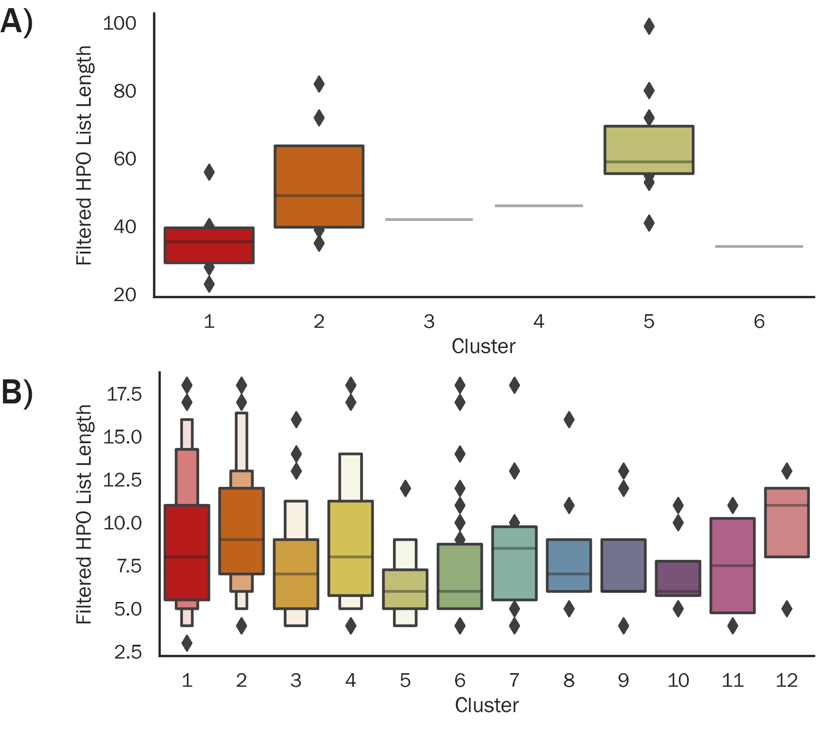

A) Box plot showing filtered HPO list for each gene group in each cluster

B) Box plot showing filtered HPO list for each patient in each cluster

**Supplementary Figure 4**

**Distribution of top-level HPO Terms in PcG and TrxG-associated conditions, by genetic diagnosis**

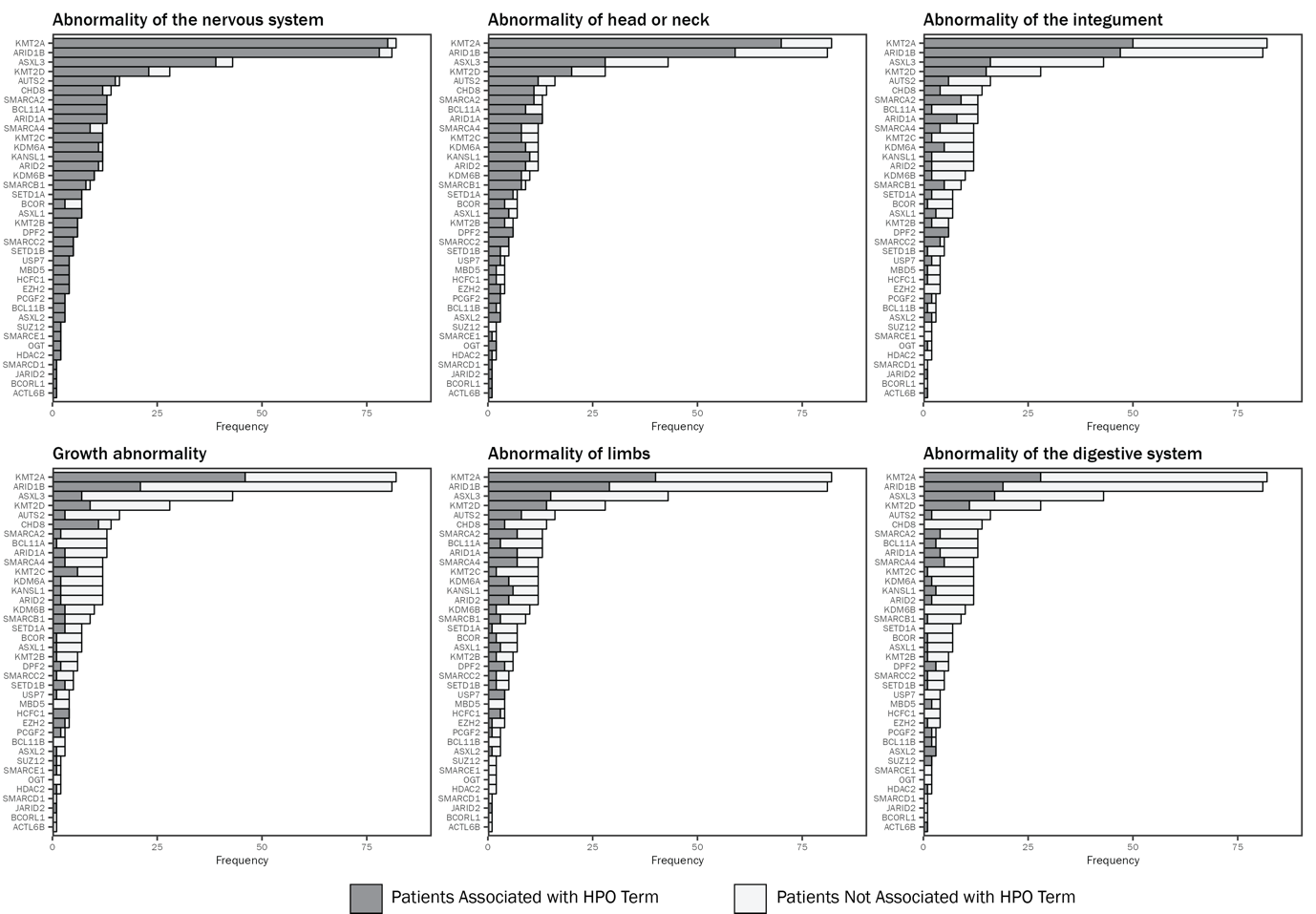

**Supplementary Figure 5**

**Distribution of patient-level cluster memberships within each gene group.**

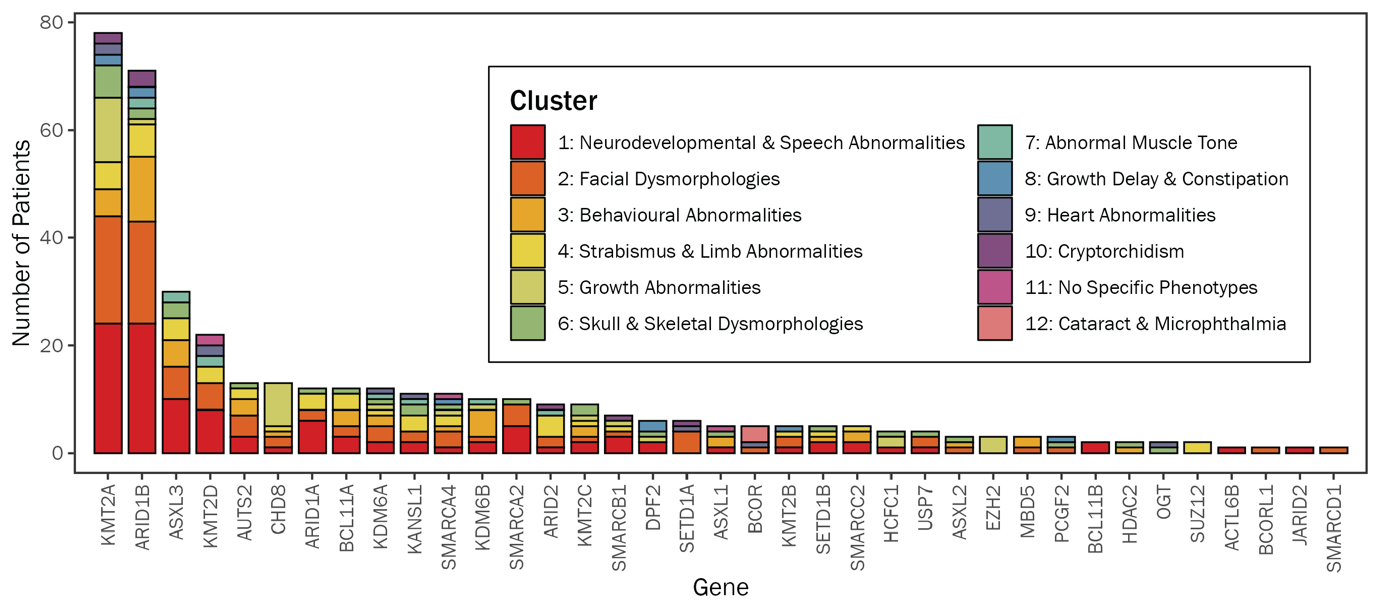
